# Correlates of Neutralizing/SARS-CoV-2-S1-binding Antibody Response with Adverse Effects and Immune Kinetics in BNT162b2-Vaccinated Individuals

**DOI:** 10.1101/2021.07.27.21261237

**Authors:** Kenji Maeda, Masayuki Amano, Yukari Uemura, Kiyoto Tsuchiya, Tomoko Matsushima, Kenta Noda, Yosuke Shimizu, Asuka Fujiwara, Yuki Takamatsu, Yasuko Ichikawa, Hidehiro Nishimura, Mari Kinoshita, Shota Matsumoto, Hiroyuki Gatanaga, Kazuhisa Yoshimura, Shin-ichi Oka, Ayako Mikami, Wataru Sugiura, Toshiyuki Sato, Tomokazu Yoshida, Shinya Shimada, Hiroaki Mitsuya

**Author notes:** Address correspondence to: Hiroaki Mitsuya or Kenji Maeda, Hiroaki Mitsuya, MD, PhD., National Center for Global Health and Medicine (NCGM) Research Institute, 1-21-1 Toyama, Shijuku, Tokyo 162-8655, JAPAN.

## Abstract

**Background:** While mRNA vaccines against SARS-CoV-2 have been exceedingly effective in preventing symptomatic viral infection, the features of immune response remain to be clarified.

**Methods:** In the present prospective observational study, 225 healthy individuals in Kumamoto General Hospital, Japan, who received two BNT162b2 doses in February 2021, were enrolled. Correlates of BNT162b2-elicited SARS-CoV-2-neutralizing activity (50% neutralization titer: NT_50_; assessed using infectious virions and live target cells) with SARS-CoV-2-S1-binding-IgG and -IgM levels, adverse effects (AEs), ages, and genders were examined. The average half-life of neutralizing activity and the average time length for the loss of detectable neutralizing activity were determined and the potency of serums against variants of concerns was also determined.

**Findings:** Significant rise in NT_50_s was seen in serums on day 28 post-1st dose. A moderate inverse correlation was seen between NT_50_s and ages, but no correlation was seen between NT_50_s and AEs. NT_50_s and IgG levels on day 28 post-1st dose and pain scores following the 2nd shot were greater in women than in men. The average half-life of neutralizing activity in the vaccinees was approximately 67.8 days and the average time length for their serums to lose the detectable neutralizing activity was 198.3 days. While serums from elite-responders (NT_50_s>1,500-fold: the top 4% among all participants’ NT_50_s) potently to moderately blocked the infectivity of variants of concerns, some serums with moderate NT_50_s failed to block the infectivity of a beta strain.

**Interpretation:** BNT162b2-elicited immune response has no significant association with AEs. BNT162b2-efficacy is likely diminished to under detection limit by 6-7 months post-1st shot. High-level neutralizing antibody-containing serums potently to moderately block the infection of SARS-CoV-2 variants; however, a few moderate-level neutralizing antibody-containing serums failed to do so. If BNT162b2-elicited immunity memory is short, an additional vaccine or other protective measures would be needed.

**Research in context:** *Evidence before this study:* While mRNA vaccines against SARS-CoV-2 have been exceedingly effective in preventing symptomatic viral infection, the salient features of immune response including the persistence of protection remain to be clarified. There is a report that anti-SARS-CoV-2 antibodies persist through 6 months after the second dose of mRNA-1273 vaccine (Doria-Rose *et al. N Engl J Med*. 2021;384:2259-2261); however, more definite immune kinetics following mRNA-vaccine-elicited protection have to be clarified. The mRNA-vaccine-elicited protection against SARS-CoV-2 variants are also to be determined.

*Added value of this study:* In the present prospective study, 225 twice-BNT162b2-dose-receiving individuals in Japan were enrolled. No significant correlation was seen between 50% neutralizing titers (NT_50_s), determined by using infectious SARS-CoV-2 virions and live target cells, and adverse effects. Largely, NT_50_s and IgG levels were greater in women than in men. Following 28 days post-2^nd^ shot, significant reduction was seen in NT_50_s, IgG, and IgM levels. The average half-life of NT_50_s was ∼68 days and the average time-length for participants’ serums to lose the detectable activity was ∼198 days. Although serums from elite-responders potently to moderately blocked the infectivity of variants of concerns, some serums with moderate NT_50_s failed to block the infectivity of a beta strain.

*Implications of all the available evidence:* BNT162b2 efficacy is likely to be diminished to under detection limit by 6-7 months post-1^st^ shot on average. Individuals with moderate NT_50_s may fail to block beta variants. If BNT162b2-elicited immune memory is lost soon, additional vaccine(s) or other protective means would be needed.

## Introduction

Since the emergence of coronavirus disease 2019 (COVID-19) caused by severe acute respiratory syndrome coronavirus 2 (SARS-CoV-2) in Wuhan, China, the disease quickly spread to the world. As of June 29, 2021, more than 180 million SARS-CoV-2-infected individuals and almost 4 million death cases have been reported in over 200 countries^1–4^. Since the beginning of the pandemic, researchers and pharmaceutical companies around the world have been working on developing vaccines^5^. Currently, more than 10 vaccines have been authorized for public use worldwide. The development of vaccines against SARS-CoV-2 was achieved time- and efficacy-wise beyond our expectations within a single calendar year from the availability of the viral sequence to the initiation of immunization of many people in several countries^6,7^.

Among various vaccines, two RNA vaccines (BNT162b2 and mRNA-1273/TAK-919) have been shown to be as much as 94-95% effective and safe^8–10^. In addition, inactivated vaccines or viral vector vaccines have also been available in certain countries and areas^5,7,9,10^. For example, the adenovirus-vector-based vaccine (ChAdOx1 nCoV-19/AZD1222) has reportedly achieved 62% efficacy in initial trials^11^. The phase 3 reports of another adenovirus-based vaccine (Ad26.COV2.S) has indicated 85% efficacy against severe disease or death^12,13^.

However, the recent emergence of various SARS-CoV-2 variants with mutations in the spike region is raising concerns about the efficacy of vaccines. The D614G and B.1.1.7 (alpha/N501Y) variants appear to be without antigenic escape^14,15^. However, the B.1.351 (beta) variant is reportedly represents a neutralization escape variant to convalescent sera^16^. The phase 3 results of NVX-CoV2373 (a nanoparticle, protein-based vaccine) from the United Kingdom indicated 89% efficacy with over 50% of cases attributable to the more transmissible alpha variant^17^. However, a phase 2b trial in South Africa showed 60% efficacy, in which approximately 90% of the endpoints occurred in subjects infected with the beta variant^18,19^, suggesting that the beta variant is less susceptible to antibodies elicited with the original Wuhan strain antigens, which is in the composition of all the vaccines currently being evaluated^7^. Another recent concern is the emergence of a B.1.617 (delta) variant, which was first detected in India, is now spreading around the world. This variant of concern (VOC) seems to have less susceptibility to vaccine-elicited protection and increased transmissibility beyond alpha strains^20^.

In the present study, we examined neutralizing activity and S1-binding-antibody response in BNT162b2-vaccinated health care workers (n=225) in Japan. We also investigated the correlation among neutralizing activity levels, S1-binding-IgG and -IgM levels, genders, and adverse events. Decline of BNT162b2-elicited immune response and activity of the elite and moderate responders against VOCs were also investigated.

## Methods

### Participants and serum specimens

Serum samples were collected from 225 vaccinated health care workers at JCHO Kumamoto General Hospital (Kumamoto, Japan). All the 225 participants were of Japanese citizen. Serum samples were analyzed at the National Center for Global Health and Medicine (NCGM) in Tokyo. The Ethics Committees from the Kumamoto General Hospital and NCGM approved this study (Kumamoto General Hospital No. 180, and NCGM-G-004176-00, respectively). Each participant provided a written informed consent, and this study abided by the Declaration of Helsinki principles. The vaccination (on days 0 and 21) and serum collection (from day 7 through day 90 post-1^st^ shot) were conducted as shown in Table 1.

**Table 1:** Study protocol and demographic characteristics of the participants.

|  |  |  |  |  |  |  |
| --- | --- | --- | --- | --- | --- | --- |
| The diagram illustrates the study protocol timeline. A horizontal timeline starts at Day 0, marked with an orange arrow labeled '1st shot'. At Day 7, a grey arrow labeled 'Test' points down. At Day 21, an orange arrow labeled '2nd shot' points down. At Day 28, a grey arrow labeled 'Test' points down. At Day 60, a grey arrow labeled 'Test' points down. At Day 90, labeled '(Post-1st shot)', a grey arrow labeled 'Test' points down. The timeline ends with an arrow pointing to the right. |  |  |  |  |  |  |
|  |  | Day7 |  | Day28 | Day60 | Day90 |
|  |  | (Post 1<sup>st</sup> dose) | (%) |  |  |  |
| All |  | 225 |  | 220 | 211 | 210 |
| Age | 20-39 | 97 | 43.1 | 92 | 84 | 84 |
|  | 40-59 | 110 | 48.9 | 110 | 109 | 108 |
|  | ≥60 | 18 | 8.0 | 18 | 18 | 18 |
|  | (Average) | (41.8 y.o.) |  |  |  |  |
| Gender | Men | 68 | 30.2 | 68 | 63 | 61 |
|  | Women | 157 | 69.8 | 52 | 148 | 149 |
| Job | Physicians | 36 | 16.0 |  |  |  |
|  | Nurses | 125 | 55.6 |  |  |  |
|  | Others | 64 | 28.4 |  |  |  |

### Cells and viruses

VeroE6_TMPRSS2_ cells^21^ were obtained from Japanese Collection of Research Bioresources (JCRB) Cell Bank (Osaka, Japan). VeroE6_TMPRSS2_ cells were maintained in DMEM supplemented with 10% FCS, 100 µg/ml of penicillin, 100 µg/ml of streptomycin, and 1 mg/mL of G418. SARS-CoV-2 NCGM-05-2N strain (SARS-CoV-2_05-2N_) was isolated from nasopharyngeal swabs of a patient with COVID-19, who was admitted to the NCGM hospital^22,23^. Five clinically isolated SARS-CoV-2 mutant strains were used in the current study: two B.1.1.7 (alpha) strains [hCoV-19/Japan/QHN001/2020 (SARS-CoV-2_QHN001_, GISAID Accession ID; EPI_ISL_804007) and hCoV-19/Japan/QK002/2020 (SARS-CoV-2_QK002_, G ISAID Accession ID; EPI_ISL_768526)] and a B.1.351 (beta) strain [hCoV-19/Japan/TY8-612-P0/2021 (SARS-CoV-2_TY8-612_)] were obtained from National Institute of Infectious Diseases, Tokyo, Japan. A B.1.617.1 (kappa) strain [TKYTK5356_2021 (SARS-CoV-2_5356_, DDBJ Accession ID; LC633761)] and a B.1.617.1 (beta) strain [hCoV-19/Japan/TKYK01734/2021 (SARS-CoV-2_1734_, GISAID Accession ID; EPI_ISL_2080609)] were provided from Tokyo Metropolitan Institute of public Health, Tokyo, Japan. Each variant was confirmed to contain each VOC-specific amino acid substitutions before the assays conducted in the present study (*vide infra*).

### Neutralization assay

The neutralizing activity of serums from vaccinated individuals was determined by quantifying the serum-mediated suppression of the cytopathic effect (CPE) of each SARS-CoV-2 strain in VeroE6^TMPRSS2^ cells as previously described with minor modifications^22^. In brief, each serum was 4-fold serially diluted in culture medium. The diluted sera were incubated with 100 50% tissue culture infectious dose (TCID_50_) of viruses at 37°C for 20 min (final serum dilution range of 1:20 to 1:4000), after which the serum-virus mixtures were inoculated to VeroE6_TMPRSS2_ cells (1.0 × 10^4^/well) in 96-well plates. For SARS-CoV-2 strains used in this assay are as follows: a wild-type strain, SARS-CoV-_205-2N_ (PANGO lineage B)^22,23^, two alpha variants (SARS-CoV-2_QHN001_ and SARS-CoV-2_QK002_), a beta variant SARS-CoV-2_TY8-612_, a delta variant SARS-CoV-2_1734_, and a kappa variant SARS-CoV-2_5336_. After culturing the cells for 3 days, the levels of CPE observed in SARS-CoV-2-exposed cells were determined using the WST-8 assay employing Cell Counting Kit-8 (Dojindo, Kumamoto, Japan). The serum dilution that gave 50% inhibition of CPE was defined as the 50% neutralization titer (NT_50_). Each serum was tested in duplicate.

### Measurement of anti-SARS-CoV-2 antibody titers

Measurement of 3 anti-SARS-CoV-2 antibody levels (anti-S1-IgG, anti-S1-IgM, and anti-N-IgG) in each participant was performed using the chemiluminescence enzyme immunoassay (CLEIA) platform (HISCL) manufactured by Sysmex Co. (Kobe, Japan) as previously reported^24^.

### Statistical analyses

Out of the 225 participants, one participant, who had been infected by SARS-CoV-2 with PCR positivity documented was primarily excluded. Demographic characteristics of the participants are described in Table 1. Correlates of neutralizing activity levels with S1-binding-IgG and -IgM levels, ages, genders, pain scores in the injection-site, and systemic fever up to 40°C were examined by Spearman rank correlation coefficient. Also, neutralizing activity levels, S1-binding-IgG, and -IgM, pain scores, systemic fever were compared between genders using the Wilcoxon rank sum test. As for participants with normal fever, their temperature was treated as 36.89 degree, a normal body temperature in Japanese^25^. Percentage of the adverse events reported in writing following the 1^st^ and 2^nd^ dose administration were determined and assessed in regard to gender. The differences in neutralization activity between each measurement time point were tested by the Wilcoxon rank sum test, and were assessed among categorized age subgroups. Similarly, difference of S1-binding-IgG and -IgM levels among time points were tested. Decline of neutralizing activity over 90 days post-1^st^ shot was assessed using the mixed-effects model including time as a main effect and intercept as a random effect. Also, the prediction slope and its 80% prediction interval were generated by drawing a sampling distribution for the fixed effects and then estimating the fitted value across that distribution. The merTools package (version 0.5.2) in R software was used for prediction. The decline for S1-binding-IgG and -IgM levels was assessed similarly. All the analyses were conducted with the use of R, version 4.1.0 (R Foundation for Statistical Computing).

## Results

### Demographic characteristics and immune response in the participants

We obtained blood samples for antibody testing from a total of 225, 220, 211, and 210 vaccine recipients on days 7, 28, 60, and 90 post-1^st^ shot, respectively (Table 1). Demographic characteristics of the participants are shown in Table 1. As of the time of enrollment, the average age of the participants was 41.8 years (range: 21 to 72 years), and 69.8% of the participants were female serving as a physician, nurse, paramedical staff, or administrative staff. None of the participants was in the immunodeficient state or was receiving immunosuppressants or steroids.

We first determined the neutralizing activity against SARS-CoV-2 in serum samples taken on day 7 post-1^st^ shot from 225 participants; however, none of the samples showed detectable neutralization activity (NT_50_ <20-fold). We then determined the neutralizing activity in samples taken on day 28 post-1^st^ shot from 220 vaccinated participants. As shown in Figure 1A, NT_50_ levels were substantially diverse among the participants: the geometric mean of NT_50_ values was 375.2 (range 25.6-2680.0). Very low or no correlation of the NT_50_ values with ages was identified (Figure 1A: Spearman’s ρ=−0.22; 95% CI −0.34 to −0.09). We also examined the levels of S1-binding-IgG and -IgM levels using the HISCL system that enables quantitative and highly sensitive determination of S1-binding-IgG and -IgM levels^24^. The geometric mean of S1-binding-IgG values was 527.0 (range 44.6-3212.2), while that of S1-binding-IgM was 85.1 (range 10.3-1406.5). There was a high positive correlation of the NT_50_ values with S1-binding-IgG levels (Spearman’s ρ=0.71; 95% CI 0.63 to 0.77) as examined on day 28 post-1^st^ shot, while there was only a low positive correlation of the NT_50_ values with S1-binding-IgM levels (Spearman’s ρ=0.43; 95% CI 0.31 to 0.53), suggesting that the neutralizing activity largely resides in IgG fraction of the serum of vaccinated participants around on day 28 post-1^st^ shot (Figures 1B and 1C). However, when examined on day 60 post-1^st^ shot, the correlations of NT_50_ levels with both IgG and IgM levels became moderate or low (Spearman’s ρ=0.56 and 0.32, respectively)(Figure S1).

**Figure 1:**
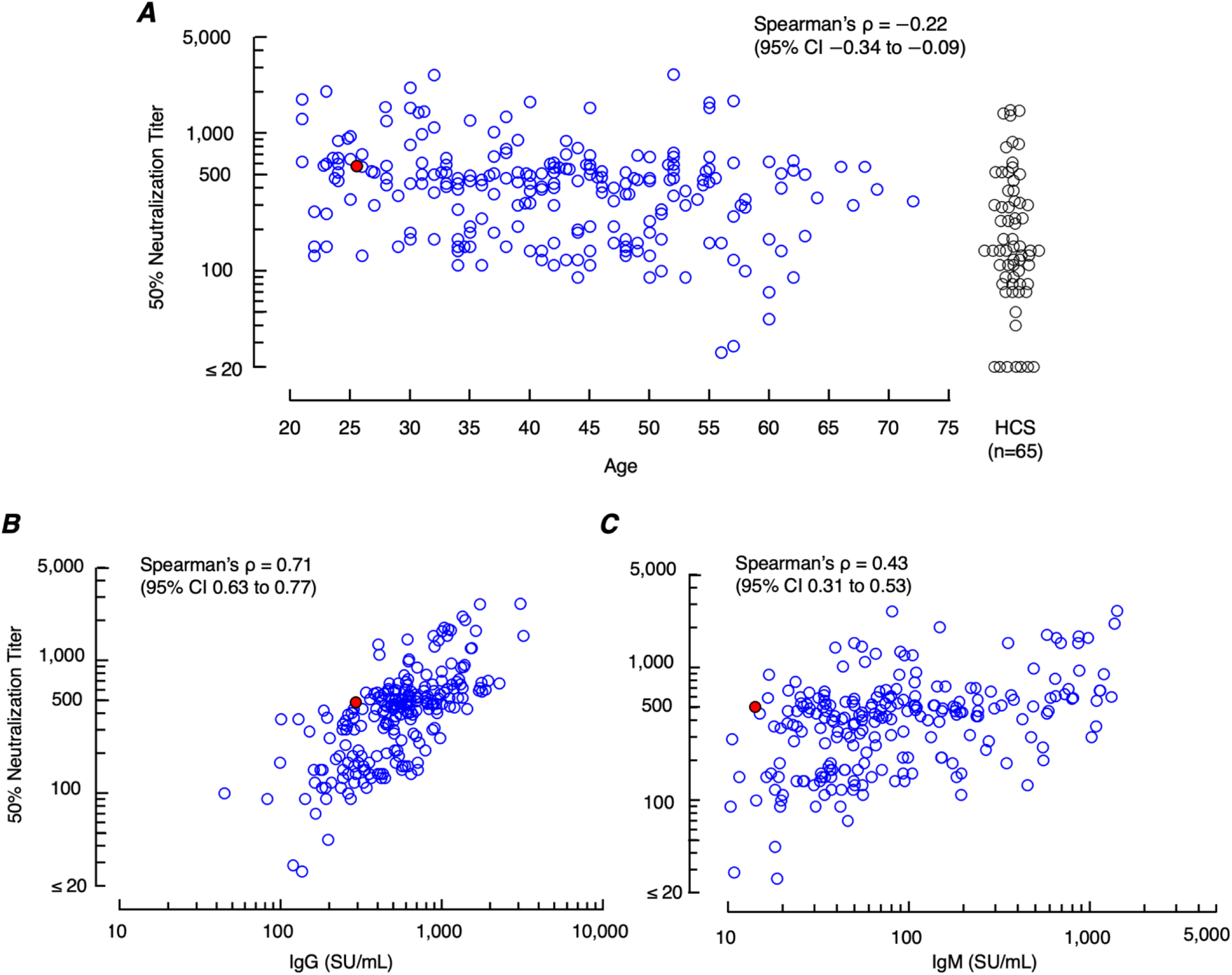
Correlations of neutralizing titers with ages and S1-binding-IgG and -IgM levels. A. Correlation between neutralizing titers (NT_50_s) and ages (on day 28 post-1^st^ shot). The age range of the study participants was 21 to 72 (average 41.8 y.o.). A correlation is negligible between NT_50_ values and ages (Spearman’s ρ=−0.22: 95% CI −0.34 to −0.09). The geometric mean NT_50_ of the values from all participants (n=225) was 375.2-fold (range 25.6-2,680-fold), greater by a factor of 2.3 than the geometric mean NT_50_ from 65 COVID-19-convalescent patients (geometric mean=163.0-fold; range 20.0-1470-fold) shown as references on the far right (human COVID-19-convalescent serum: HCS). B. A high correlation is identified (Spearman’s ρ=0.71; 95% CI 0.63 to 0.77) between NT_50_ values and S1-binding-IgG levels in samples obtained on day 28 post-1^st^ dose. C. Moderate correlation is seen between neutralizing titers and S1-binding-IgM levels (Spearman’s ρ=0.43; 95% CI 0.31 to 0.53). One participant, who had been infected with SARS-CoV-2 with PCR-positivity documented, is indicated as a solid-red solitary circle. This participant was excluded from all analyses at later timepoints.

### The occurrence of adverse effects has no association with the BNT162b2-elicited neutralizing activity levels

Commonly observed adverse events reported following BNT162b2 vaccination include injection-site pain, systemic fever, headache, and fatigue^10^. In the present study, the events were observed largely more often following the 2^nd^ shot (Figure S2) as previously reported by Polack *et al*.^10^ Pains in the inject-site were reported by 67.6 and 61.6% of the participants and systemic fever (≥37.1°C) was reported by 3.6 and 46.4% of the participants following the 1^st^ and 2^nd^ shots, respectively. Since the severity of pains can be relatively more quantitatively rated than that of other adverse effects such as headache and fatigue, the possible correlate of the NT_50_ values with the severity of pains rated with the short form McGill pain questionnaire^26^ was first examined. No correlation was seen between the NT_50_ values and the pain grades assessed following the 2^nd^ shot (Spearman’s ρ=0.14; 95% CI 0.00 to 0.26). The correlation was also negligible between the NT_50_ values and the incidence of systemic fever (Spearman’s ρ=0.26; 95% CI 0.13 to 0.38)(Figures 2A and 2B).

**Figure 2:**
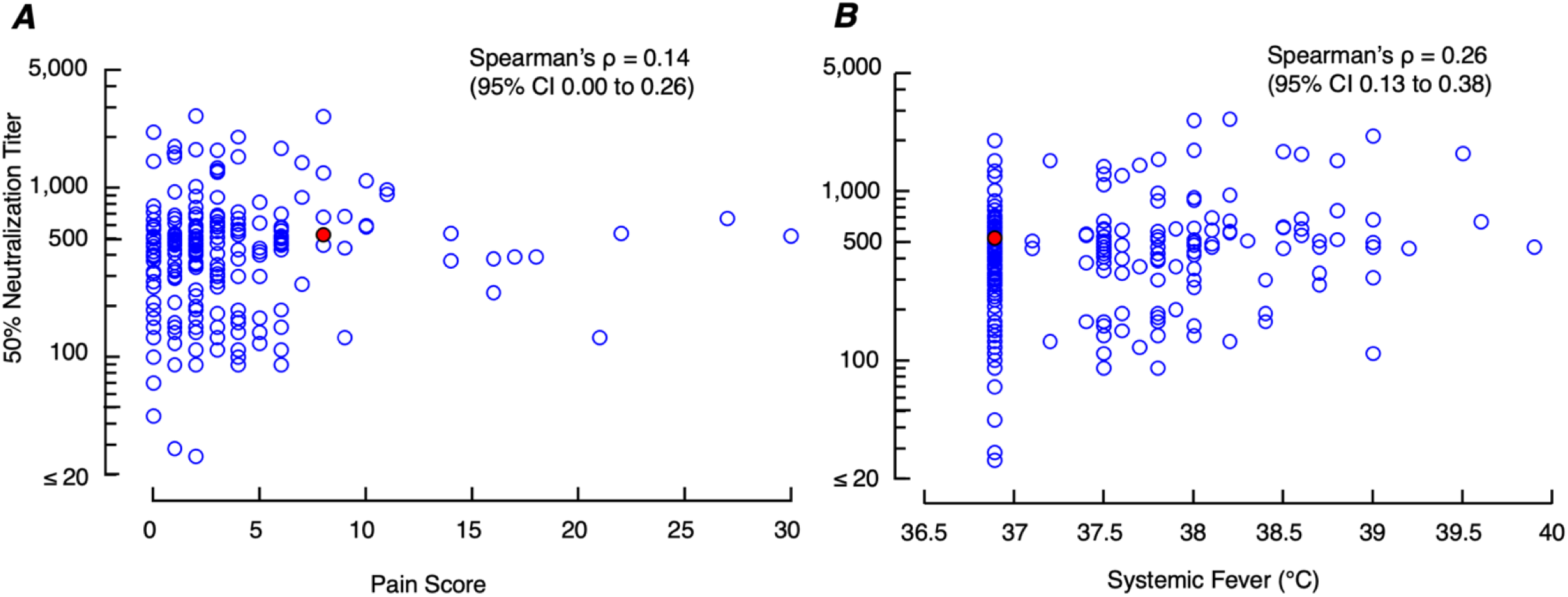
Correlations of neutralizing titers with injection-site pain scores and systemic fever grades. A. No correlation was seen between NT_50_ values and injection-site pain (Spearman’s ρ=0.14; 95% CI 0.00 to 0.26). The injection-site pain following the 2^nd^ BNT162b2 dose was scored by using the short-form McGill Pain Questionnaire^26^. B. Correlation was negligible between NT_50_ values and systemic fever grades (Spearman’s ρ=0.26; 95% CI 0.13 to 0.38). A solid-red circle indicates a person with previous SARS-CoV-2 infection documented.

### The average half-life of neutralizing activity in the vaccinees is approximately 67.8 days and the average time-length for their serums to lose the detectable activity is 198.3 days

Considering that recent multiple clinical studies strongly suggest that the presence of high-level neutralizing antibodies is generally sufficient to confer protection against SARS-CoV-2 infection and that the protection against COVID-19 development is largely explained by robust SARS-CoV-02-neutralizing antibody response^8–10^. If so, the once-established neutralizing antibody levels will decrease in time. We thus examined at what rate the levels of NT_50_ and S1-bindng-IgG and -IgM levels change by determining those levels from the data on day 28 (n=220), day 60 (n=211), and day 90 (n=210) post-1^st^ shot (Figures 3A-C). The reduction of all NT_50_, IgG, and IgM levels from day 28 through day 90 post-1^st^ shot was found to occur virtually linearly. By computation, the predicted average half-life of all the NT_50_ values turned out to be 67.8 days and those of S1-binding-IgG and IgM levels were 53.5 days and 43.6 days, respectively (Figure 3D). The half-life of the NT_50_ values and that of S1-binding-IgG were reasonably comparable, corroborating that the neutralizing activity largely resides in the S1-binding IgG fraction. Based on the chronologically linear nature of the reduction identified, we attempted to extrapolate from such half-life values and tried to predict the average time-length for the serums of the participants having significant NT_50_ values to lose the activity down to under the undetectable level (UDL)(*<*20-fold)(Figure 3D). The predicted average time-length for the serums to lose the activity was computated to be 198.3 days, while that of the top 10% participants to lose the activity was 204.3 days. The time-length of the middle 10% (between the top 45% and 55%) participants to lose the activity was 187.6 days. For all participants, it was predicted that day 160 after the 1st shot was when the 80% lower limit of predicted NT_50_ levels drops under the detection level (UDL), while day 237 after the 1st shot was when the 80% upper limit of predicted NT_50_ levels drops below UDL. Similarly, for the top 10% participants, the estimated days dropping below UDL were 171 and 243 for lower and upper limits, respectively, while for the middle 10%, the estimated days were 155 and 224, respectively. As for IgG, the predicted time-length for the serums to get undetected was 346.8 days, and days reaching below UDL were 333 and 362 for lower and upper limits, respectively. Likewise, the estimated day reaching below UDL for IgM was 183.6, and the lower and upper limits were 170 and 198 days, respectively (Figure 3D). We also asked whether the chronological decay rate of neutralization titers and S1-binding-IgG and -IgM differs among three age subgroups: (i) 20-39 yo, (ii) 40-59 yo, and (iii) 60’s and beyond. No significant difference was identified among the three age subgroups in the levels of neutralizing titers, IgG, or IgM levels (*p*=0.60, 0.16, and 0.11, respectively: Figures S3A-C). The present data suggest that vaccinated individuals with good neutralization response would lose BNT162b2’s protection in 6 to 7 months without regard to age subgroups unless such people achieve robust immune boost response upon the future exposure to SARS-CoV-2. Otherwise, they should be protected by another booster vaccine shot or by other protective means.

**Figure 3:**
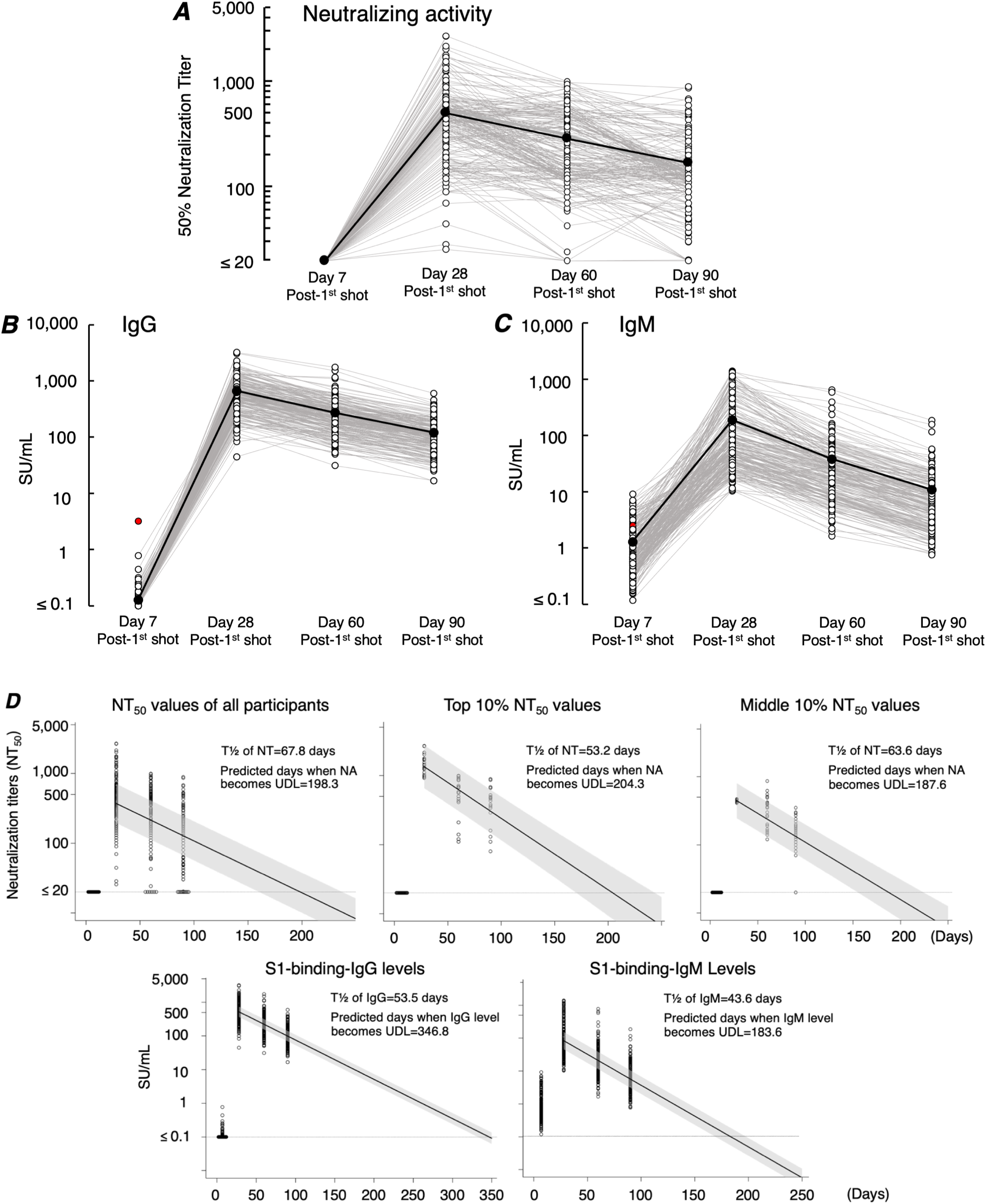
Kinetics of neutralizing activity and S1-binding-IgG and -IgM levels. Time-course analyses of neutralizing activity for 90 days were conducted. The 1^st^ vaccine was administered on day 0, and the 2^nd^ vaccine on day 21. Blood samples from vaccinated individuals were obtained on days 7, 28, 60, and 90 post 1^st^ shot as illustrated in Table 1. A. Neutralizing activity is shown as NT_50_ (50% neutralizing titer). The NT_50_ value of 20-fold is the detection limit and values determined to be less than 20-fold were treated as 20-fold. B and C. Kinetics of S1-binding-IgG and -IgM levels are shown. The average values of each data point are shown in black solid circles, which are connected with solid black lines. One participant, who had been infected with SARS-CoV-2 with PCR-positivity documented, is indicated as a solid-red solitary circle in B and C. This participant was excluded from all analyses at later timepoints. D. Decline of neutralizing activity, S1-binding-IgG and -IgM over 90 days post-1^st^ shot. The solid-black lines consist of predicted values estimated by mixed effects model, and the shaded areas denote corresponding 80% prediction intervals. The dashed horizontal lines in the upper three panels denote the NT_50_ detection limit of 20-fold. NT_50_ values determined to be less than 20-fold were treated as 20-fold. The lowest detection limit for S1-binding-IgG and -IgM quantification shown as dashed horizontal lines in the two lower panels was 0.1 SU/ml and the values lower than 0.1 SU/ml were calculated as 0.1 SU/ml.

### Neutralization titers, S1-binding-IgG levels, and pain scores in the injection site were greater in women than in men

We then asked whether there were differences between genders in neutralization activity levels, S1-bindng-IgG and -IgM levels, injection-site pain scores, and systemic fever grades. Statistically significant differences were identified in the levels of neutralization determined on 60 and 90 days post-1^st^ shot (p=0.002 and 0.002, respectively), S1-binding-IgG levels determined on 28, 60, and 90 days (p<0.001, p=0.001, and p=<0.001, respectively) post-1^st^-shot, and S1-binding-IgM levels on 60 and 90 days post-1^st^ shot (p=0.025 and 0.044, respectively)(Figures S4A-C). The injection-site pain score was greater in women (p<0.001) (Figure S4D). However, there was no difference in systemic fever grades between genders (Figure S4E). However, no difference was seen in the decline rates of neutralization activity, S1-binding-IgG and -IgM levels between men and women (Figure S5).

### Some serums retain potent activity against various VOCs, but others showed substantially less potent or undetectable activity

We finally asked whether the neutralizing antibodies elicited by BNT162b2 vaccination blocked the infectivity and replication of various variants of concerns (VOCs). To this end, we employed serum samples from 6 elite responders (NT_50_ values >1,500-fold: the top 4% of all participants’ NT_50_ values as determined on 28 days post-1^st^ dose) and serum samples from twelve randomly-selected moderate responders (NT_50_ values=200∼1,500-fold) and tested them for their inhibition of the infectivity and cytopathic effect of each variant in the VeroE6_TMPRSS2_ cell-based assay^21^. As shown in Figure 4, serums from the elite responders (n=6) showed potent inhibition against SARS-CoV-2_05-2N_ (Wuhan strain, PANGO lineage B), while they showed less activity against SARS-CoV-2_QHN001_ and SARS-CoV-2_QK002_ (alpha), SARS-CoV-2_5356_ (kappa), SARS-CoV-2_1734_ (delta), and SARS-CoV-2_TY8-612_ (beta).

**Figure 4:**
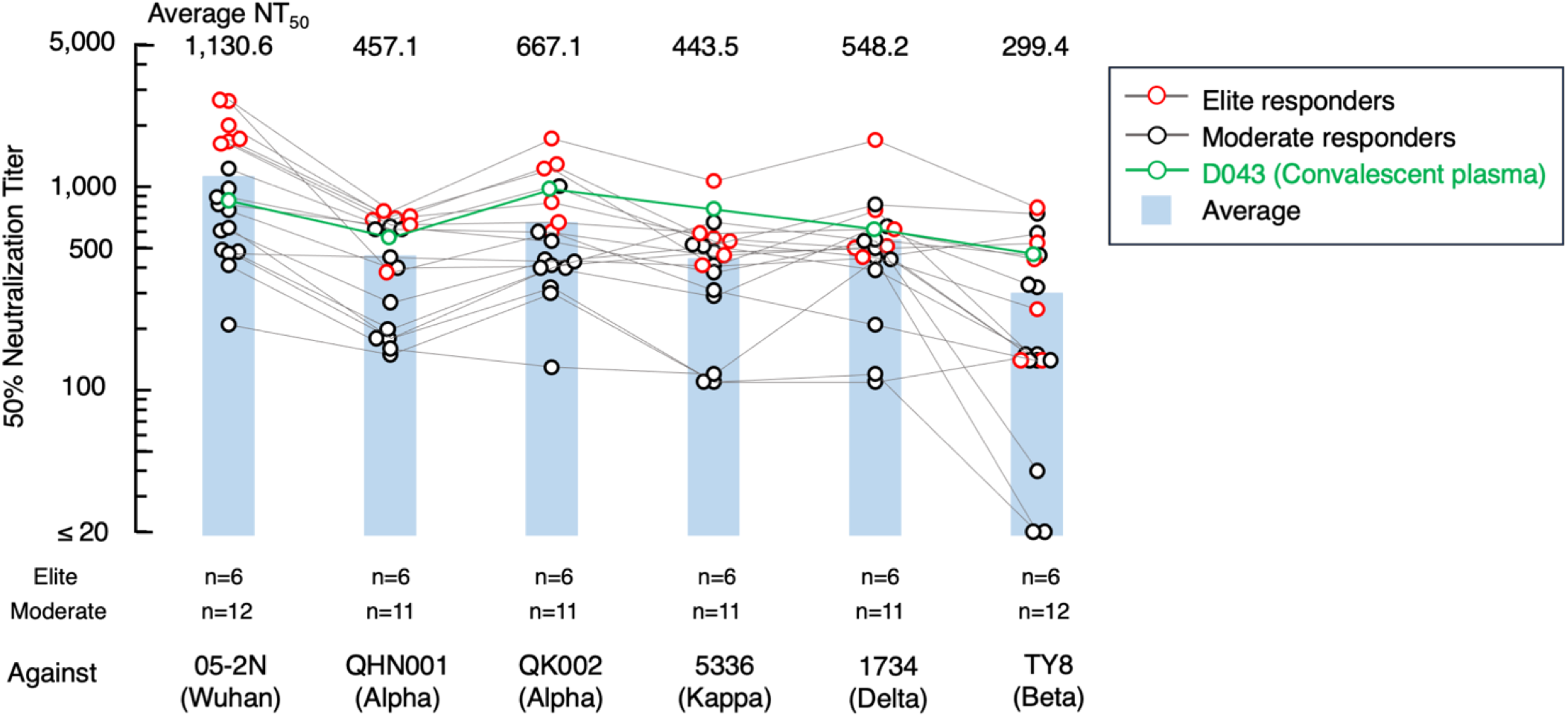
Blockade of the infectivity and replication of SARS-CoV-2 variants by vaccinees’ serums. The activity of vaccinees’ serums to block the infectivity and replication of 5 SARS-CoV-2 variants (alpha variants: SARS-CoV-2_QHN001_ and SARS-CoV-2_QK002_; a beta strain: SARS-CoV-2_TY8-612_; a delta strain: SARS-CoV-2_1734_; and a kappa strain: SARS-CoV-2_5356_) was evaluated. A Wuhan strain 05-2N^19^ was employed as a reference SARS-CoV-2. Six serums were from elite responders (NT_50_ >1,500-fold) and 12 serums were from randomly-selected moderate responders (NT_50_=200∼1,500-fold). The NT_50_ titers of each serum against 6 SARS-CoV-2 strains are shown in red circles (for 6 elite responders) and in black circles (for 12 moderate responders). D043 is a serum from a COVID-19-convalescent patient^39^ and served as an internal control in the assays.

Serums from moderate responders (n=12) exerted less activity against the Wuhan strain than those from the elite responders. Some serums from the moderate responders also showed substantially low potency to all the VOCs tested. Notably, three serums from the moderate responders showed only marginal activity against SARS-CoV-2_TY8-612_ (beta). Two of those three samples had no detectable inhibitory activity against SARS-CoV-2_TY8-612_ (Figure 4).

## Discussion

In this prospective observational study, 225 healthy individuals [physicians (n=36), nurses (n=125), and other healthcare professionals (n=64)], who received two doses of 30 µg BNT162b2 (Pfizer–BioNTech) vaccine in February 2021, were enrolled, and the correlates of neutralization activity represented by 50% neutralization titers (NT_50_) determined by employing the target living VeroE6^TMPRSS2^ cells and live SARS-CoV-2 with ages, adverse effects (AEs) that occur often such as injection-site pain and systemic fever were examined. The kinetics of NT_50_ values and S1-binding antibody levels were also examined. There was a significant rise in the NT_50_ values as determined on day 28 post-1^st^ shot (a week after post 2^nd^ shot) compared to those on day 7 post-1^st^ shot. Correlation was negligible between NT_50_ values and ages or systemic fever grades. In this regard, most adverse effects that occur within 1-3 days following vaccine shots are thought to be caused by the release of certain pyrogenic and inflammatory cytokines (*e*.*g*., interleukin-1, interleukin-6, and tumor-necrosis factor) from antigen-presenting cells (APCs) such as macrophages and dendritic cells when they ingest and process the exogenous antigens (*i*.*e*., SARS-CoV-2 spike antigens) and transmit the antigenic information to relevant immune cells. Such early-phase defense events include response to antigenic determinants irrelevant to neutralizing activity but those eliciting S1-binding antibodies. The release of the pyrogenic and inflammatory cytokines mostly subsides within days following the vaccine shot. The released cytokines activate the antigen-specific-antibody-producing B-cells, which respond to the processed antigenic determinants presented by the APCs and start to produce antigen-specific antibodies such as neutralizing antibodies as well as non-neutralizing but S1-binding antibodies. Such antigen-specifically-activated and antibody-producing B-cells continue to produce antibodies. In the case of BNT162b2 vaccination, it appears that it takes 10 to 12 days from the 1^st^ vaccine shot for the vaccinated individuals to achieve the amounts of neutralizing antibodies that are enough to block the infection of substantial numbers of the virally-targeted cells and to inhibit further spread of the infection^9,10^. It is thought that the release of pyrogenic and inflammatory cytokines and the build-up of the protective antibody levels are different events, occurring chronologically ∼10-12 days apart. These two different events appear to have resulted in the absence of significant correlates between NT_50_ levels and AEs examined in the present study.

In the present study, the NT_50_ values had a substantial correlation with S1-binding-IgG levels but had only moderate correlation with S1-binding IgM levels, suggesting that the major neutralizing activity resides within the S1-binding IgG fraction. Interestingly, the approximate half-life of NT_50_ values (67.8 days) and that of S1-binding-IgG levels (53.5 days) were reasonably close to each other, corroborating the assumption of the presence of the major fraction of neutralizing antibodies within IgG fraction. In human body, IgG has concentration-dependent half-life of approximately 21 days and IgM around 5-6 days^27^. By contrast, the half-lifes of NT_50_, S1-bindng-IgG, and -IgM levels determined in the present study were much longer with 43.6-67.8 days. This discrepancy is perhaps attributed to the persistence of continuously-antibody-producing B cells over weeks or months in the body of the participants following the vaccination^28,29^, thereby the half-lifes of neutralizing activity and S1-binding-IgG and -IgM levels have been extended as compared to the physiological half-lifes of IgG and IgM. However, the assumption of the half-lifes in the present work was based on the chronologically linear nature of the reduction observed during the present study period (Figure 3D). Also, the sensitivity and quantitativeness of S1-binding-IgG and -IgM levels determined with using the chemiluminescence enzyme immunoassay (CLEIA) platform (HISCL)^24^ were much greater (the dynamic range is 0.1 to 2,000 SU/ml)^24^ than that of neutralizing activity, whose dynamic range is 20 to 4,000^22^. Thus, the time length of S1-binding-IgG becoming under detection levels was calculated to be longer than that of neutralizing activity, although it should be noted that the protective effect of BNT162b2 judged by neutralizing activity is most likely associated with clinical outcomes. In addition, recently, Doria-Rose et al. reported that anti-SARS-CoV-2 antibodies persist through 6 months after the second dose of mRNA-1273 administration^30^. However, the study was of a relatively small scale (n=33), and more definite data are needed for constructing more protective measures. However, a caution should be used in assuming half-lifes since we presently have no knowledge as to how long neutralizing antibody- or S1-binding antibody-producing B-cells continue to produce antibodies following the administration of two doses of BNT162b2. If such B-cells produce antibodies for a shorter period of time than we assumed in the present study, the half-lifes of neutralizing and S1-binding antibodies could be shorter than we estimated in the present work.

There is a growing body of evidence that COVID-19 results in more severe symptoms and greater mortality among men than among women^31,32^. A cohort study of 17 million adults in England has revealed a strong association between male sex and the risk of death from COVID-19 (hazard ratio 1.59, 95% confidence interval 1.53–1.65)^33^. In the present data set, significantly greater levels of NT_50_, S1-binding-IgG and -IgM were documented in women than in men when examined on 28, 60, and 90 days post-1^st^ shot, while there was no difference in either of NT_50_, S1-binding-IgG or -IgM levels on day 7 post-1^st^ dose (Figures S4A-C). These results apparently relate to the findings by others reporting that women, in general, have more robust ability to control infectious pathogens (*i*.*e*., SARS-CoV-2) than men^33,34^. Indeed, there is increasing evidence indicating strong correlation between SARS-CoV-2-neuralizing antibody titers and clinical efficacy, suggesting that a neutralizing antibody response provides the primary contribution to protection against COVID-19^35^ and that the presence of high levels of neutralizing antibody is largely sufficient for protection against SARS-CoV-2 infection and clinical onset upon exposure to the virus^36,37^. In fact, Imai *et al*. have reported that the administration of convalescent plasma from previously-SARS-CoV-2-infected hamsters completely protected newly SARS-CoV-2-exposed hamsters from contracting viral pneumonitis^38^. Thus, the greater neutralizing activity in women than in men observed in the present study can contribute at least in part to the gender differences in COVID-19 disease outcomes. Also, of note, the number of participants with ages greater than 60 years was rather small (18 of 225), which might have made the statistical power insufficient to find significant differences with other two age groups (20-39 yo and 40-59 yo groups)(Figures S3A-C).

We also examined how the BNT162b2-elicited neutralizing antibodies blocked the infectivity and cytopathic effect of five different variants of concerns in the cell-based assays using various infectious variants (one Wuhan strain, two alpha strains, one strain each of beta, delta and kappa strains). Six selected serums from elite responders showed quite potent activity to the alpha, kappa, and delta variants, while they exerted relatively moderate activity against the beta strain (Figure 4). On the other hand, one of the randomly-selected 12 serums from moderate responders showed a marginal activity (NT_50_ value of 40-fold) and two of them failed to show detectable activity (NT_50_ values <20-fold) against the beta strain (Figure 4). These data suggest that BNT162b2-receiving vaccinees who develop high magnitudes of neutralizing antibody should probably be well protected against the infection by most variants; however, those who develop only low levels of neutralizing antibody may be vulnerable to the infection by certain variants such as beta strains. If so, to such low-responders to BNT162b2 even after the 2^nd^ shot, an additional 3^rd^ shot may be needed. If the 3^rd^ dose of the same vaccine fails to elicit good levels of neutralizing antibodies, new types of vaccines with different platform have to be stratified.

## Data Availability

All data presented in this paper are included in the manuscript.

## Contributors

KM and HM had access to all data in this study and took and hold all responsibility for the integrity of the data and the accuracy of the data analysis. KM and HM: Concept and design. KM, MA, KT, TM, KN, YT, HG, KY, SO, TS, and TY: Acquisition, analysis, or interpretation of data. YU, YS, AF, and HM: Statistical analysis. AF, YI, HN, MK, SM, AM, WS, and SS: Administrative, technical, or material support. KM and HM: Original draft writing. All authors: Reading and writing manuscript.

## Acknowledgments

This research was supported in part by a grant from the Japan Agency for Medical Research and Development to Maeda (grant number JP20fk0108260, 20fk0108502) and to Mitsuya (grant number 20fk0108502), and in part by a grant for MHLW Research on Emerging and Re-emerging Infectious Diseases and Immunization Program to Maeda (grant number JPMH20HA1006) from Ministry of Health, Labor and Welfare, and in part by a grant for COVID-19 to Mitsuya (grant number 19A3001) and Maeda (grant number 20A2003D) from the Intramural Research Program of National Center for Global Health and Medicine, and in part by the Intramural Research Program of the Center for Cancer Research, National Cancer Institute, National Institutes of Health (Mitsuya). The authors thank Drs. Kenji Sadamasu, Mami Nagashima, Hiroyuki Asakura, and Mr. Isao Yoshida for providing two SARS-CoV-2 variants, Drs. Nobuyo Higashi-Kuwata, Shinichiro Hattori, Kouki Matsuda, Ms. Mariko Kato and Ms. Sachiko Otsu for helping some experiments, and Dr. Norihiro Kokudo for critical discussion for the manuscript.

## Supplementary appendix

**Figure S1:**
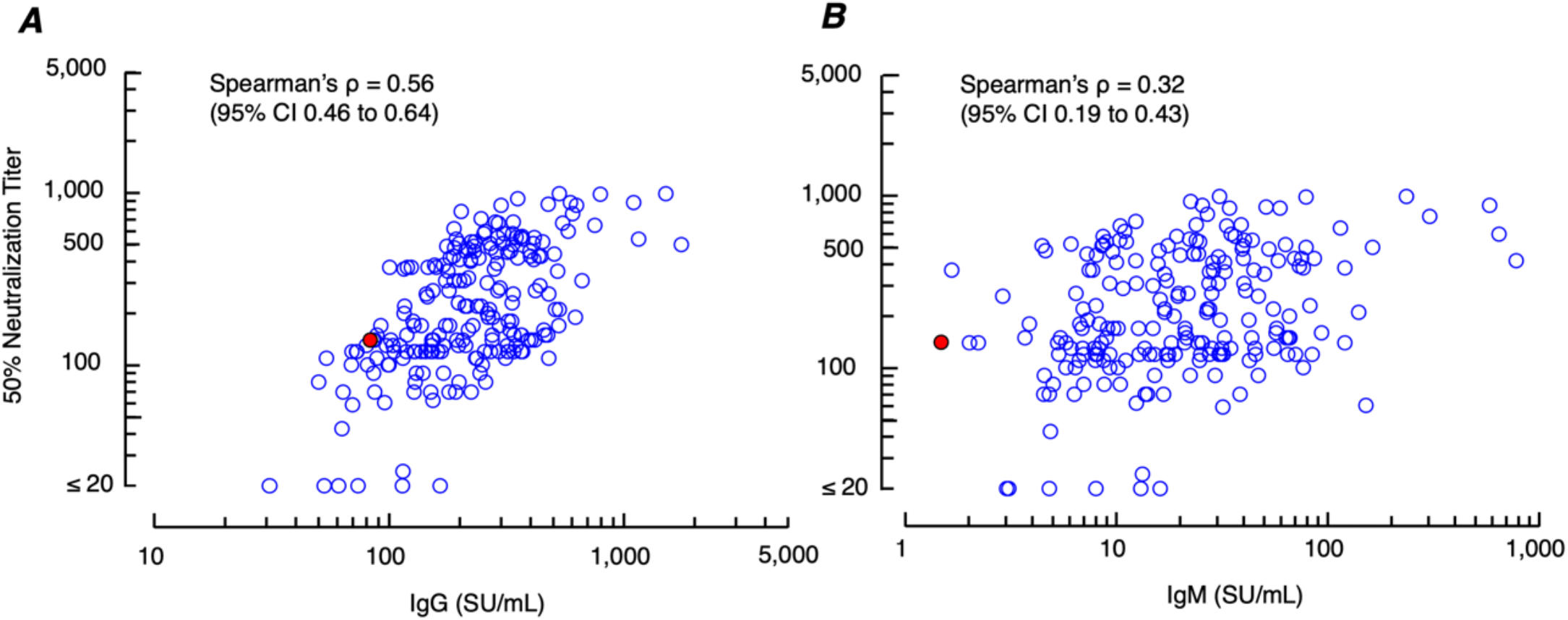
A. Correlation between neutralizing activity and IgG levels on day 60 post 1^st^ shot. Moderate correlation is identified (Spearman’s ρ=0.56; 95% CI 0.46 to 0.64) between NT_50_ values and S1-binding-IgG levels in samples obtained on day 60 post-1^st^ dose. B. Correlation between neutralizing activity and S1-binding-IgM levels on day 60 post-1^st^ dose. Low correlation is seen between neutralizing titers and S1-binding-IgM levels (Spearman’s ρ=0.32; 95% CI 0.19 to 0.43). A red-solid circle denotes a person with previous SARS-CoV-2 infection documented.

**Figure S2:**
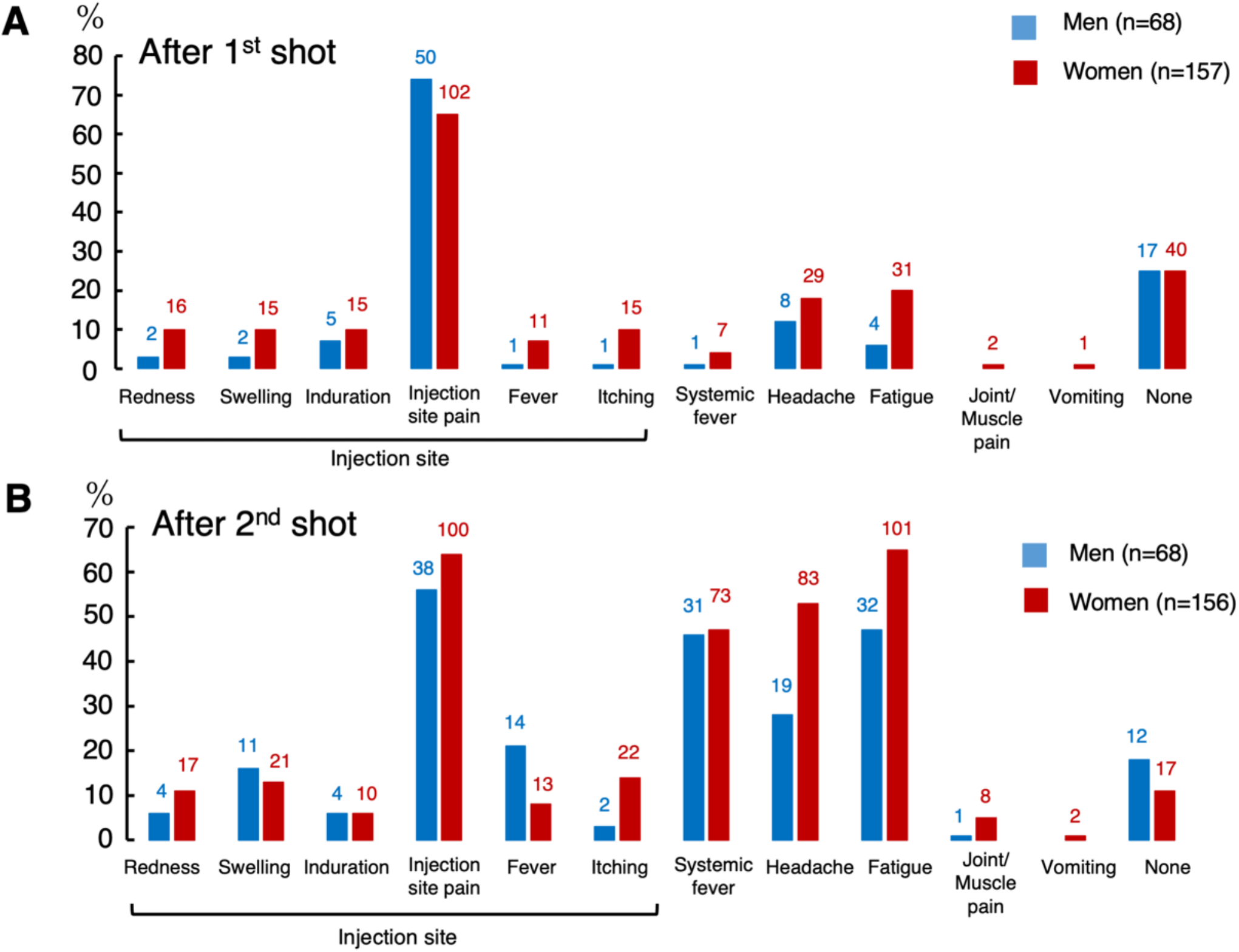
Incidences of adverse effects reported after the 1^st^ vaccination (A) and the 2^nd^ vaccination (B). A total of 225 (Men: 68, Women: 157) participants reported after the 1^st^ shot, and a total of 224 (Men: 68, Women: 156) participants after the 2^nd^ shot. Systemic fever of ≥37.1°C and pain scores of ≥1 were taken into the analyses. The number at the top of each bar denotes the number of individuals reporting each AE.

**Figure S3:**
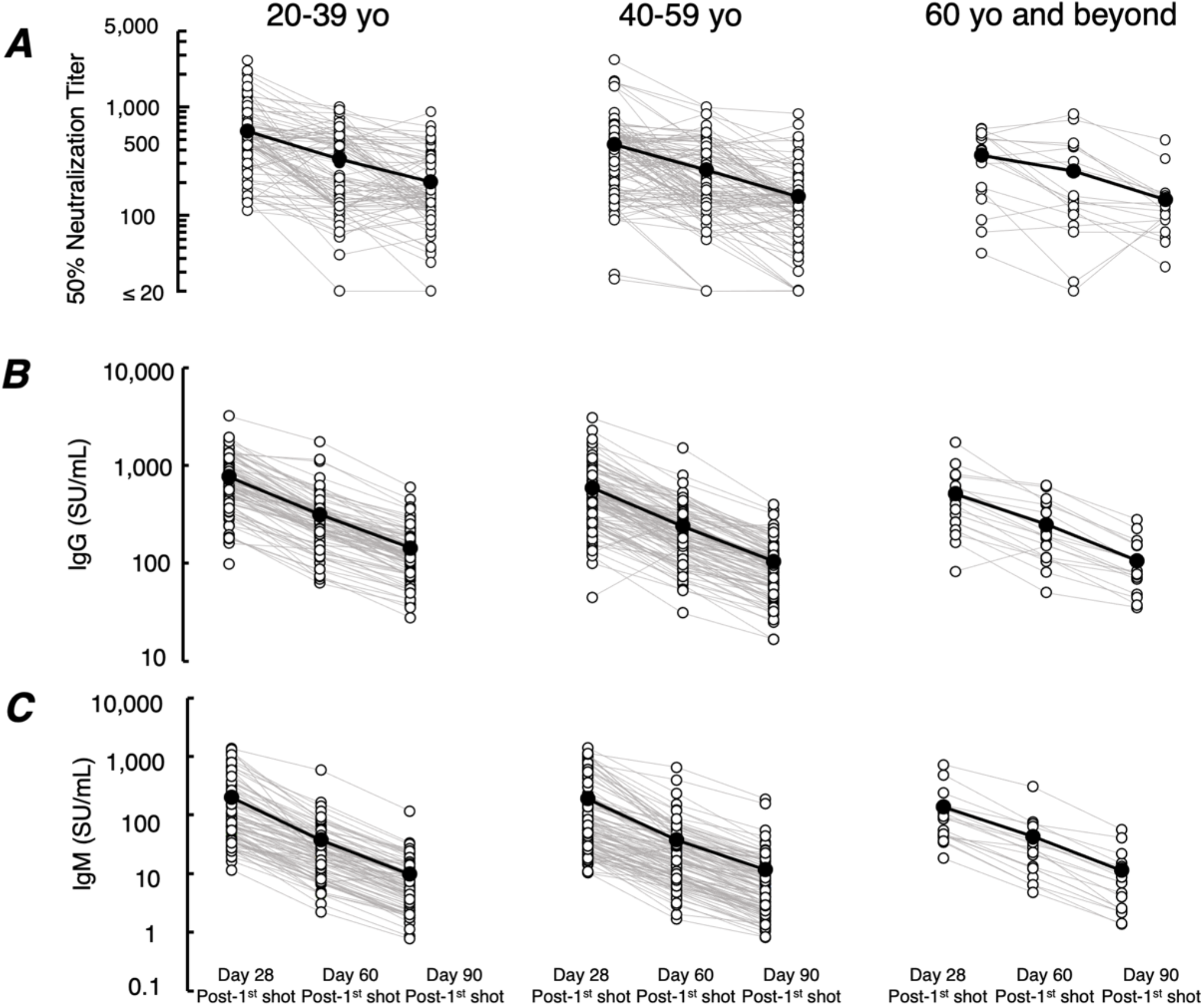
Decline of neutralization titers and S1-binding antibody levels in three age groups. The chronological decline rates of neutralization titers (A), S1-binding-IgG (B), and -IgM levels (C) among three age groups; (i) 20-39 yo, (ii) 40-59 yo, and (iii) 60’s and beyond, were evaluated. No significant difference was identified among the three age groups in the decline rates of neutralizing titers, IgG, or IgM levels (*p*=0.596, 0.163, and 0.106, respectively). Statistical analysis was conducted with the mixed-effects model including time, age category and time-age category interaction term and intercept as a random effect. A solitary open circle on day 28 in the 20-39 yo subgroup illustrated in B denotes a participant who did not provide blood sample for unknown reason on days 60 and 90 post-1^st^ shot.

**Figure S4:**
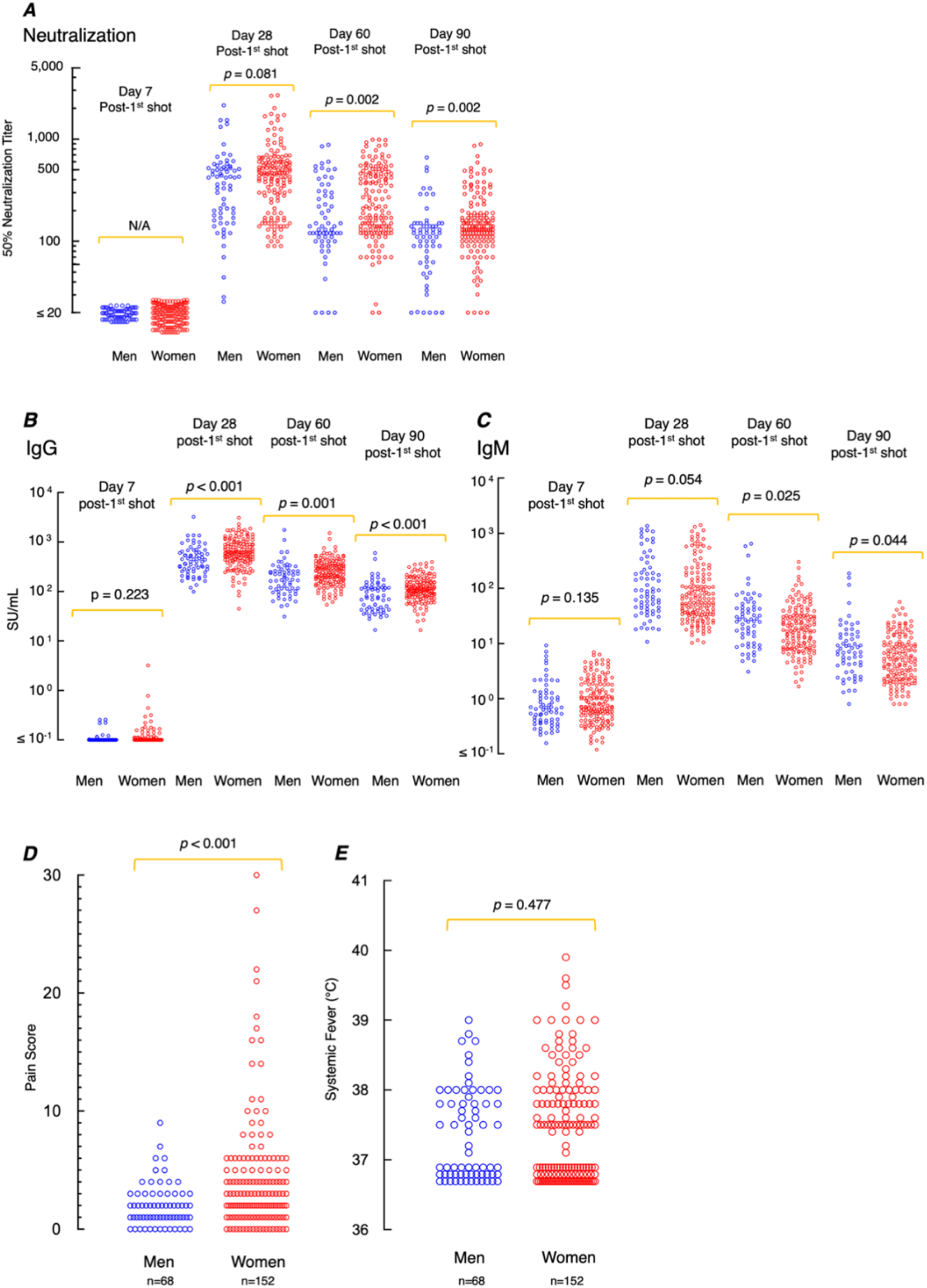
Neutralization activity, S1-binding-IgG and -IgM levels, and pain scores are greater in women than in men. A. Neutralization activity was greater in women than in men at two timepoints (days 60 and 90 post-1^st^ shot). B and C. S1-binding-IgG levels were greater in women at all three timepoints (days 28, 60, and 90 post-1^st^ shot) and S1-binding-IgM levels greater in women at two timepoints (day 60 and 90 post-1^st^ shot). D, E. Injection-site pain scores and systemic fever grades in men and women. Scores of injection-site pain were greater in women than in men (*p*<0.001)(D), while no difference was seen in systemic fever grades between the two groups (*p*=0.477)(E). Statistical significance was evaluated using Wilcoxon rank sum test. N.A., not applicable.

**Figure S5:**
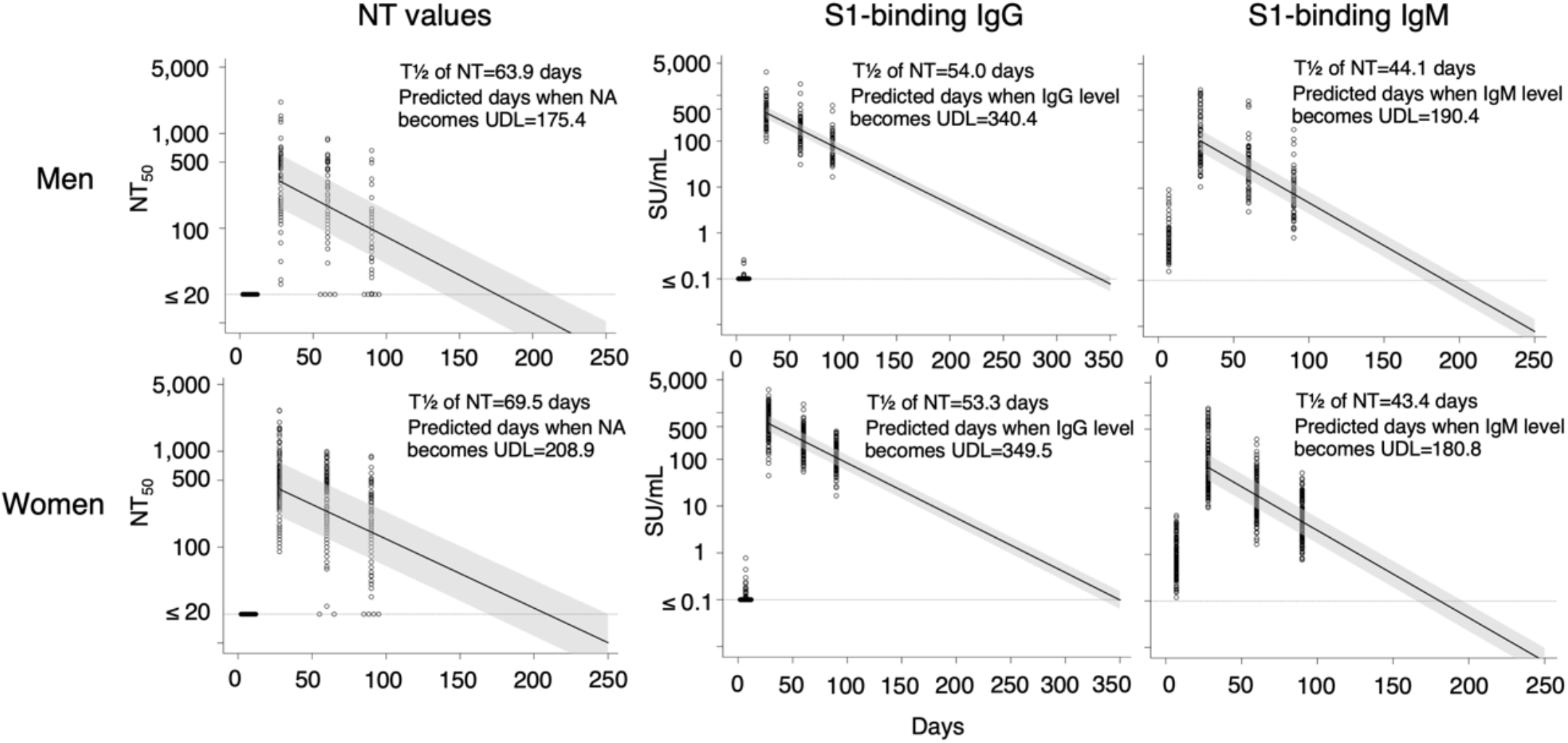
No difference in the decline rates of the neutralization activity, S1-binding-IgG and -IgM levels between men and women. Decline rates of neutralizing activity, S1-binding-IgG and -IgM over 90 days post-1^st^ shot were compared. The solid lines consist of predicted values estimated by mixed effects model, and the shaded areas denote corresponding 80% prediction intervals. The dashed horizontal lines denote the NT_50_ detection limit (20-fold). NT_50_ values determined to be less than 20-fold were treated as 20-fold. The lowest detection limit for S1-binding-IgG and -IgM quantification was 0.1 SU/ml and the values lower than 0.1 SU/ml were calculated as 0.1 SU/ml. No significant difference was identified in the three indicators between men and women. Statistical analysis was conducted with the mixed-effects model including time, age category and time-age category interaction term and intercept as a random effect.

